# Polygenic contributions to suicidal thoughts and behaviors in a sample ascertained for alcohol use disorders

**DOI:** 10.1101/2022.08.18.22278943

**Authors:** Sarah MC Colbert, Niamh Mullins, Grace Chan, Jacquelyn L Meyers, Jessica Schulman, Samuel Kuperman, Dongbing Lai, John Nurnberger, Martin H Plawecki, Chella Kamarajan, Andrey P Anokhin, Kathleen K Bucholz, Victor Hesselbrock, Howard J Edenberg, John Kramer, Danielle M Dick, Bernice Porjesz, Arpana Agrawal, Emma C Johnson

## Abstract

Suicidal thoughts and behaviors have partially distinct genetic etiologies. We used PRS-CS to create polygenic risk scores (PRS) from GWAS of non-suicidal self-injury, broad sense self-harm ideation, non-fatal suicide attempt, death by suicide, and depression. Using mixed-effect models, we estimated whether these PRS were associated with a range of suicidal thoughts and behaviors in the Collaborative Study on the Genetics of Alcoholism (N = 7,526). All PRS were significantly associated with suicidal ideation and suicide attempt (betas=0.08-0.44, FDR<0.023). All PRS except non-suicidal self-injury PRS were associated with active suicidal ideation and severity of suicidality (betas=0.04-0.22, FDR<0.034). Several associations remained significant in models where all significant PRS were included as simultaneous predictors, and when all PRS predicted suicide attempt, the PRS together explained 6.2% of the variance in suicide attempt. Significant associations were also observed between some PRS and persistent suicidal ideation, non-suicidal self-injury, attempt severity and desire to die. Our findings suggest that PRS for depression does not explain the entirety of the variance in suicidal thoughts and behaviors, with PRS specifically for suicidal thoughts and behaviors making additional and sometimes unique contributions.

## INTRODUCTION

While several risk factors for suicidality are known [1–5], little research has identified which factors may be particularly important to the progression from suicidal ideation to suicide attempt to death by suicide [6–8]. Currently, the best predictors of suicide death are the preceding stages. For example, high level of intent (measured using Beck’s Suicidal Intention Scale [9]) at previous attempt and having made a medically serious previous attempt were found to be strong predictors of suicide death [10,11]. However, longitudinal studies following individuals with a prior non-fatal suicide attempt found that a modest proportion (1.2%-6.7%) die by suicide [12–14]. In contrast to death by suicide, suicide attempt is well predicted by its risk factors: 29% of those reporting suicidal ideation and 56% of those reporting ideation and a plan, also endorse suicide attempts [15].

Genome-wide association studies (GWAS) of broad sense self-harm ideation (i.e., thoughts of self-harm regardless of suicidal intent) [16,17], self-harm [16,17], suicide attempt [18] and death by suicide [19] have revealed significant SNP-heritability (the proportion of phenotypic variance that is explained by all SNPs included in a GWAS) estimates (h^2^_SNP_=0.068-0.25). Non-suicidal self-injury and suicidal thoughts and behaviors are transdiagnostic, co-occurring with psychiatric disorders ranging from bipolar disorder [20] to eating disorders [21], but are most frequently studied in the context of depression. Polygenic risk scores (PRS; an estimate of one’s genetic liability to a trait of interest) for major depressive disorder, depressive symptoms and/or broad depression are associated with self-reported suicidal thoughts and behaviors ranging from self-harm (in general) to suicidal ideation, suicide attempt and death by suicide [19,22–27]. However, major depressive disorder [28] is only partially genetically correlated with suicidal ideation (r_g_=0.46) [17] and suicide attempt (r_g_=0.78) [18] and depression [29] is also not perfectly genetically correlated with death by suicide (r_g_=0.42) [30]. Some studies have found significant associations between observed suicidal thoughts and behaviors and PRS of self-harm [16,23] and suicide attempt [18,24,31]. However, with the exception of a recent study from Mullins *et al*. [18] which identified a significant association between suicide attempt and a PRS constructed from a GWAS of suicide attempt conditioned on major depressive disorder [18], little research has explored whether genetic liability to non-suicidal self-injury and suicidal thoughts and behaviors that are independent from genetic liability to depression explain variance in suicidal thoughts and behaviors. Thus, validating the transdiagnostic heritability of various aspects of suicidal thoughts and behaviors requires that PRS constructed from GWAS of non-suicidal self-injury and suicidal thoughts and behaviors be examined in other samples to estimate whether this polygenic liability is an independent predictor of suicidal thoughts and behaviors, beyond the genetics of depression. Such independent effects would further solidify the need to consider the genetics of suicidal thoughts and behaviors as transdiagnostic markers of liability to a broad range of psychopathology with suicidal thoughts and behaviors as components.

Non-suicidal self-injury and suicidal thoughts and behaviors may represent a genetically heterogeneous set of phenotypes. However, genetic research to date has struggled to investigate this question due to the common overlap amongst these phenotypes. Twin and family studies have been best able to answer these questions using distinct phenotypes; for example, a recent study from Kendler *et al*. [32] found evidence to suggest that suicide attempt and death by suicide are genetically distinct in ways beyond severity. Still, some relationships between these phenotypes are less clear, with some twin studies having found that a large portion of the variation in suicide attempt is explained by genetic factors shared with non-suicidal self-injury [33] and that the two behaviors share a genetic correlation of 0.94 [34], while a study from Russell *et al*. [35] was not able to identify any genetic correlation between non-suicidal self-injury and suicide attempt, although in a considerably smaller sample. Ability to explore these genetic relationships using GWAS data has been much more limited by omnibus phenotype definitions. For example, GWAS of suicide attempt frequently include cases who have died by suicide [25], and GWAS of self-harm behavior often do not specify intent, consequently combining non-suicidal self-injury and suicide attempt into one measure [16,17]. The inconsistent findings regarding genetic similarities and distinctions between suicidal thoughts and behaviors necessitate further exploration using current, well-powered GWAS data.

Using GWAS of non-suicidal self-injury, broad sense self-harm ideation [36], non-fatal suicide attempt [18], death by suicide [19] and depression [29,37], we examined whether polygenic liability to these phenotypes was associated with 11 suicidal thoughts and behaviors in individuals in the Collaborative Study on the Genetics of Alcoholism (COGA). As COGA was primarily ascertained for alcohol use disorders (AUD), individuals in the sample may have elevated risk for other mental health conditions including suicidal thoughts and behaviors, and its independent assessment of these suicidal thoughts and behaviors apart from depression supports the use of COGA to address these questions. We hypothesized that: (1) PRS for suicidal thoughts and behaviors would explain additional, unique variance in suicidal thoughts and behaviors, beyond that explained by depression PRS and (2) non-suicidal self-injury would be genetically distinct from suicidal thoughts and behaviors.

## METHODS

### DATA

#### Target Dataset

COGA was designed to investigate the genetic underpinnings of AUD and has collected data from extended families with an extensive AUD history as well as comparison families from the community [38–40]. Using the Semi-Structured Assessment for the Genetics of Alcoholism (SSAGA) interview [41,42], COGA collected data from all participants, on a wide variety of mental health phenotypes, including depression and, independently, suicidal thoughts and behaviors. Many questions about suicidal thoughts and behaviors were nested, meaning that they were only asked as follow-up questions to individuals endorsing a pre-requisite variable. For example, data for active suicidal ideation was only collected for individuals who endorsed any suicidal ideation. The nine mental health phenotypes used in the current study are:

- Non-suicidal self-injury: Individuals were asked, “(Other than when you tried to take your own life,) did you ever hurt yourself on purpose, for example, by cutting or burning yourself?”
- Suicidal ideation: Individuals were asked, “Have you ever thought about killing yourself?”
- Active suicidal ideation: Individuals endorsing suicidal ideation were subsequently asked “Did you have a plan? (Did you actually consider a way to take your life?)”
- Persistent suicidal ideation: Individuals endorsing suicidal ideation were subsequently asked “Did those thoughts persist for at least 7 days in a row?”
- Suicide attempt: Individuals were asked “Have you ever tried to kill yourself?”
- More than one attempt: Individuals who endorsed a suicide attempt were asked “How many times (did you try to kill yourself)?”. Affected individuals answered more than once.
- Medical treatment after attempt: Individuals who endorsed a suicide attempt were asked “Did you require medical treatment after you tried to kill yourself?”
- Substance use prior to attempt: Individuals who endorsed a suicide attempt were asked “Did you try to kill yourself after you had been drinking?” and “Did you try to kill yourself after using drugs?”. Affected individuals answered yes to either one or both of these questions.
- Desire to die: Individuals who endorsed a suicide attempt were asked “Did you really want to die?”. Affected individuals answered “maybe” or “yes” to this question. It may seem inconsistent that an individual would endorse having attempted suicide and answer no to this question, however suicidal desire and suicidal intent may actually be separate facets of suicidality [43], with only the latter being required to classify self-injurious behavior as an attempt. Considering this, individuals answering this question retrospectively may have responded no if they experienced regret during their attempt or upon reflection realized they may not have truly wanted to die despite intending to die.

In addition to the above SSAGA defined phenotypes, we constructed a binary variable (“attempt severity”) which compared individuals with a suicide attempt and at least one co-occurring behavior considered to make it more severe (more than one attempt, requiring medical treatment after an attempt, substance use prior to an attempt, desire to die) to individuals endorsing only a single suicide attempt with no co-occurring conditions. We also created an ordinal variable measuring “severity of suicidality”. To do so, we first created a measure in which individuals were given a score of 0 to 4, with increasing scores reflecting greater severity, i.e., no suicidal thoughts and behaviors= 0, non-suicidal self-injury= 1, suicidal ideation= 2, persistent or active suicidal ideation= 3, and suicide attempt= 4. For individuals with multiple assessments, an assignment was made using the highest endorsed category. A description of all binary phenotypes and their prevalence in the sample can be found in **Table 1**.

As discovery GWAS (see below) were only available in samples of European ancestry, and accuracy of PRS prediction is low when genetic ancestries of discovery and target datasets differ, our analyses were limited to individuals of European genetic ancestry. Genetic data and analysis covariates were available for 7,646 individuals of European ancestry. The data processing has been described in more detail previously [44]. Because the COGA sample was genotyped using multiple arrays, QC focused on a common set of 47,000 high quality variants that were genotyped in all arrays, and duplicate individuals were removed. Briefly, variants were imputed separately for each array using the Haplotype Reference Consortium panel and were filtered to only include SNPs with INFO scores > 0.8, minor allele frequency > 0.01 and that passed Hardy-Weinberg equilibrium (HWE p<10^−6^).

### Discovery Datasets

We used summary statistics from existing GWAS of broad sense self-harm ideation, non-fatal suicide attempt, death by suicide and depression and constructed our own GWAS of non-suicidal self-injury to calculate PRS for individuals in the COGA sample.

- Broad sense self-harm ideation: GWAS summary statistics (N= 135,819) were derived from European individuals in the UK Biobank who answered either “*No*”, “*Yes, once*”, or “*Yes, more than once*” to “*Have you contemplated harming yourself (for example by cutting, biting, hitting yourself or taking an overdose)?*” (Data-Field 20485, and was analyzed as an ordinal variable, i.e., “No”= 0, “Yes, once”= 1 and “Yes, more than once”= 2). We refer to this form of thinking as broad sense self-harm ideation as suicidal intent could not be determined. The summary statistics were downloaded from the Pan-UKB GWAS results (https://pan.ukbb.broadinstitute.org).
- Non-fatal suicide attempt: Suicide attempt summary statistics came from a meta-analysis of GWAS of suicide attempt by the International Suicide Genetics Consortium (ISGC) using a combination of clinical interviews, self-report questionnaires, hospital records and ICD-10 codes [18]. Suicide attempt cases were categorized as individuals who made a non-fatal suicide attempt (an act of self-harm performed with intent to die) or died by suicide (∼20% of cases). As there is prior evidence to suggest partly distinct genetic etiologies of suicide attempt and death by suicide [45], we obtained summary statistics from a GWAS of non-fatal suicide attempt only (i.e., excluding individuals who had died by suicide) from the ISGC (N= 507,744, N case= 23,767) in order to keep these phenotypes independent.
- Death by suicide: GWAS summary statistics (N= 18,223, N case= 3,413) were taken from data from the Utah Office of the Medical Examiner [19].
- Non-suicidal self-injury: Previous GWAS have not specifically looked at non-suicidal self-injury, disentangling it from suicide attempt, so we conducted our own GWAS for non-suicidal self-injury in the UK Biobank sample using SAIGE [46]. A direct measure of non-suicidal self-injury is not provided by the UK Biobank; however, we derived this phenotype using information from the UK Biobank’s on-line “Thoughts and Feelings” mental health questionnaire. The questionnaire asked individuals “*Have you deliberately harmed yourself, whether or not you meant to end your life?*” (Data-Field 20480, N = 137,969), and those who answered “Yes” (N = 5,924) were then asked in a follow-up question, “*Have you harmed yourself with the intention to end your life?*” (Data-Field 20483). Individuals who answered yes to both questions (N = 3,056), indicating self-harm with suicidal intent, were excluded from our analyses. After quality control (see below), we were left with a remaining sample size of 133,620 individuals (N cases = 2,845). SAIGE performs a GWAS in two steps: first, it uses individual level genotype data to fit the null logistic model, then uses imputed dosage data to perform the association tests [46]. Step 1 of SAIGE used genotype array data (i.e., non-imputed) which was filtered to exclude variants with > 5% missingness, Hardy-Weinberg equilibrium p-values below 1e-10 and minor allele frequency < 0.05. Following QC, genotype array data underwent LD pruning in PLINK (using the command --indep-pairwise 50 5 0.2). Step 2 of SAIGE was performed using imputed dosages downloaded from the UK Biobank filtered for INFO score > 0.9, minor allele frequency > 0.01 and minor allele count > 20. We limited our UK Biobank sample to individuals of European genetic ancestry using the ancestry assignments published by the Pan-UKB team [36]. Following the procedure from Zhou et al. (2018), we included sex, age and the first 10 genetic principal components (provided by the Pan-UKB team) as covariates in the GWAS. We implemented the leave-one-chromosome-out option in SAIGE. The GWAS produced no genome-wide significant (p < 5e-8) results; the strongest association (p = 9.21e-7) was at lead SNP rs72778956 on chromosome 10 (Figure S1, Table S1).
- Depression: We used METAL [47] to meta-analyze two GWAS of depression (N= 750,414, N case= 254,566): one from the Million Veteran Program sample [37] and the other from a previous meta-analysis of the UK Biobank and Psychiatric Genomics Consortium [29].

## ANALYSES

### Polygenic risk scores

PRS in COGA individuals were calculated with PRS-CS [48], using effect sizes from the previously mentioned GWAS summary statistics. For depression and non-fatal suicide attempt, we allowed the software to learn the global shrinkage parameter from the data. However, when calculating PRS using the non-suicidal self-injury, broad sense self-harm ideation and death by suicide GWAS, for which sample sizes were more limited, we manually set the global shrinkage parameter phi=1e-2 (following guidance from the PRS-CS development team and others [48,49]). We used PLINK’s --score command [50] to produce individual level PRS in the COGA sample. PRS were scaled to a standard normal distribution.

### Association Models

We used logistic mixed-effect regression models to formally test for associations between the non-suicidal self-injury, broad sense self-harm ideation, non-fatal suicide attempt, death by suicide and depression PRS and suicidal thoughts and behaviors in COGA. As an exception, we used a linear mixed-effect regression model to test for associations between PRS and the ordinal severity of suicidality variable we constructed. All models included sex, age at last interview, genotyping array, and the first 10 genetic principal components as fixed covariates and family ID as a random intercept. Due to the ascertainment strategy for COGA families, we also tested whether significant associations between the PRS and suicidal thoughts and behaviors remained after additionally including AUD as a covariate. We used the MuMIn [51] package in R to calculate pseudo-R^2^ (percent variance explained) values.

For suicidal thoughts and behaviors that were associated with multiple PRS, we constructed follow-up “multi-PRS” models which accounted for those PRS as predictors simultaneously. For all models, we considered false discovery rate (FDR) < 0.05 as significant. FDR was calculated separately in each set of models, i.e., across the main single PRS models, then across the models covarying for AUD, then across the multi-PRS models.

### Average PRS Visualization

To visualize the mean PRS values for affected and unaffected individuals, we fit a mixed effects model to get PRS residual values by regressing the following covariates on the PRS: sex, age at last interview, genotyping array, the first 10 genetic principal components, and family ID (as a random effect). For each suicidal thought or behavior, we split individuals into “affected” and “unaffected” groups and then calculated the average PRS residuals and standard errors in each of the groups for visualization purposes (shown in Fig. 1). As this figure compares affected and unaffected groups, only original binary phenotypes from COGA are included in the figure (i.e., derived phenotypes including attempt severity and severity of suicidality are excluded).

**Figure 1.**
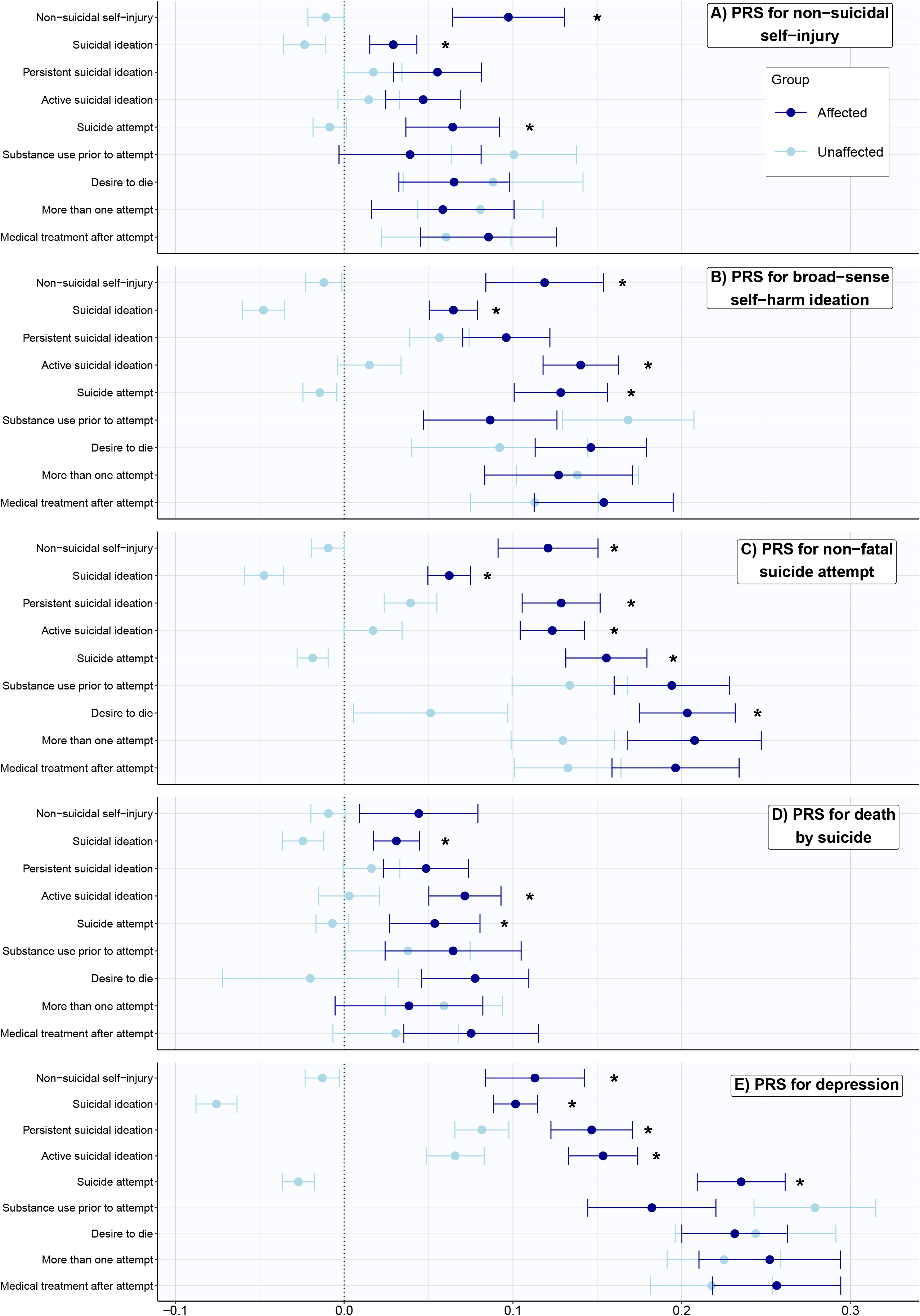
Average PRS for individuals exhibiting specific Suicidal thoughts and behaviors. PRS studied were non-suicidal self-injury, self-harm ideation, suicide attempt, death by suicide and depression. Dots represent average PRS residuals after regressing out covariates and bars represent standard errors. Dark blue dots represent average PRS in the affected groups, while light blue dots represent average PRS in the unaffected groups. Asterisks indicate associations that are significant (see Table 2). A) average PRS residuals for non-suicidal self-injury, B) average PRS residuals for self-harm ideation, C) average PRS residuals for non-fatal suicide attempt, D) average PRS residuals for death by suicide, E) average PRS residuals depression.

### Genetic correlations

We calculated genome-wide genetic correlations (r_g_) among the five GWAS using LD score regression [52].

## RESULTS

To maximize the total sample size in each model, our analytic sample included individuals who had complete genetic data, information for all covariates and data for at least one suicidal thought or behavior (e.g., individuals with suicide attempt data but not non-suicidal self-injury data could still be included in the models with suicide attempt as an outcome). A total of 6,434 individuals had data on non-suicidal self-injury (prevalence= 0.09; see Table 1). 7,526 individuals had data on suicidal ideation (prevalence= 0.43) and suicide attempt (prevalence= 0.11). Of those self-reporting suicidal ideation, 41% indicated active suicidal ideation and 31% indicated persistent ideation. In the group of individuals self-reporting suicide attempt, 40% had endorsed more than one attempt. Those who self-reported a suicide attempt were asked follow-up questions about their attempt, or if they had endorsed multiple attempts were asked follow-up questions in regards to their most serious attempt. Of these individuals who reported at least one suicide attempt, 44% required medical treatment after their attempt, 45% had used a substance prior to their attempt and 72% had indicated desire to die.

PRS were correlated with each other to varying degrees (see Table S2), with the highest correlation between non-fatal suicide attempt and depression (r = 0.29) and the lowest correlation between non-suicidal self-injury and death by suicide (r = 0.02). Overall, we found that average PRS residual values were higher in individuals endorsing suicidal thoughts or behaviors compared to their counterparts who did not endorse suicidal thoughts or behaviors (shown in Fig. 1), although when formally testing for associations between PRS and observed suicidal thoughts and behaviors using logistic mixed-effect regression models, not all associations were significant after FDR corrections (shown in Table 2).

We found that all PRS were significantly associated with suicidal ideation (beta= 0.08-0.31, se= 0.026-0.028, FDR < 0.004, R^2^ = 0.002-0.025) (shown in Table 2). Given that all PRS were significantly associated with suicidal ideation in separate models, we constructed a multi-PRS model in which all five PRS simultaneously predicted suicidal ideation. In the multi-PRS model, the depression, non-fatal suicide attempt and broad sense self-harm ideation PRS remained significantly associated with suicidal ideation (FDR < 1.10e-3), while the death by suicide PRS and non-suicidal self-injury PRS were no longer significantly associated with suicidal ideation (shown in Table S3).

Similarly, we observed that all PRS were significantly associated with suicide attempt (beta= 0.10-0.44, se= 0.040-0.044, FDR< 0.023, R^2^ = 0.003-0.047), but the strongest association was with non-fatal suicide attempt PRS and the weakest association was with the non-suicidal self-injury PRS. In the follow-up multi-PRS model which included all PRS as simultaneous predictors (R^2^ = 0.062), associations only remained significant between suicide attempt and the depression and non-fatal suicide attempt PRS.

Active suicidal ideation was significantly associated with all PRS (beta= 0.14-0.22, se= 0.039-0.043, FDR< 0.003, R^2^ = 0.005-0.011) except for the non-suicidal self-injury PRS, and associations with all PRS remained significant, although they were slightly weaker, in the multi-PRS model (beta= 0.11-0.16, se= 0.041-0.045, FDR < 0.024, R^2^ = 0.023). We also observed significant, although comparatively weak, associations between severity of suicidality and all PRS (beta= 0.04-0.10, se= 0.015-0.016, FDR< 0.034, R^2^ = 0.002-0.013) except for the non-suicidal self-injury PRS. In the multi-PRS model in which all PRS, except for the non-suicidal self-injury PRS, simultaneously predicted severity of suicidality (R^2^ = 0.021), only the depression and non-fatal suicide attempt PRS remained significant predictors (shown in Table S3).

Persistent suicidal ideation was significantly associated with the depression PRS (beta= 0.12, se= 0.042, FDR= 0.016, R^2^ = 0.003) and non-fatal suicide attempt PRS (beta= 0.18, se= 0.044, FDR= 2.2e-4, R^2^ = 0.007). However, in the multi-PRS model, the association between persistent suicidal ideation and the depression PRS was no longer significant, while the association with the non-fatal suicide attempt PRS remained significant (beta= 0.16, se= 0.05, FDR= 0.002). Desire to die was only significantly associated with non-fatal suicide attempt PRS (beta= 0.26, se= 0.098, FDR= 0.019, R^2^ = 0.009); similarly, attempt severity was only associated with the non-fatal suicide attempt PRS (beta= 0.33, se= 0.15, FDR= 0.049, R^2^ = 0.017).

Non-suicidal self-injury was significantly associated with all PRS (beta= 0.16-0.24, se= 0.049-0.053, FDR< 0.0035, R^2^ = 0.005-0.011) except for the death by suicide PRS. Notably, in the multi-PRS model in which non-suicidal self-injury was predicted by all PRS except for the death by suicide PRS, associations with the PRS remained significant with the exception of non-suicidal self-injury’s association with the non-suicidal self-injury PRS. More than one attempt, requiring medical treatment after an attempt and substance use prior to an attempt were not significantly associated with any of the PRS (shown in Table 2).

Additionally, we tested whether PRS remained significantly associated with suicidal thoughts and behaviors after controlling for AUD by adding it as an additional covariate in the models. We found that all previously significant associations (shown in Table 2) remained significantly associated (FDR < 0.05) in the models covarying for AUD (shown in Table S4).

Finally, we calculated significant, positive genetic correlations amongst all five GWAS (r_g_ = 0.54-0.84, all p < 6e-4; shown in Table S5). Death by suicide and suicide attempt were most strongly genetically correlated (r_g_ = 0.84, se = 0.08, p = 7.1e-24), while the weakest genetic correlation was observed between death by suicide and self-harm ideation (r_g_ = 0.53, se = 0.11, p = 9.2e-7). In general, non-suicidal self-injury had the weakest genetic correlations with the other traits (r_g_ = 0.54-0.69, se = 0.09-0.16, all p < 6e-4).

## DISCUSSION

We explored the associations of polygenic liability for non-suicidal self-injury, broad sense self-harm ideation, non-fatal suicide attempt, death by suicide, and depression with a variety of suicidal thoughts and behaviors observed in the COGA sample, a family-based sample that was enriched for AUD diagnoses. All PRS were significantly associated with suicidal ideation and suicide attempt, and active suicidal ideation and severity of suicidality were associated with all PRS except for the non-suicidal self-injury PRS. In models where depression and suicidal thoughts and behaviors were included as simultaneous predictors, many associations remained significant, supporting our hypothesis that while genetic liability to suicidal thoughts and behaviors and depression are related, they retain some independent genetic contributions to suicidal thoughts and behaviors. On the other hand, non-suicidal self-injury PRS did not remain significantly associated with any self-harm behaviors in COGA after accounting for the depression and suicidal thoughts and behaviors PRS, suggesting that beyond its shared genetics with the other PRS, non-suicidal self-injury PRS is not significantly associated with suicidal thoughts and behaviors in this sample. Additionally, in models testing the association between single PRS and active suicidal ideation and severity of suicidality, non-suicidal self-injury PRS was the only PRS that was not significantly associated, further suggesting that non-suicidal self-injury is somewhat genetically distinct from suicidal thoughts and behaviors. A follow-up exploratory analysis in which we calculated genetic correlations between the non-suicidal self-injury and suicidal thoughts and behaviors GWAS provided partial support for a genetic distinction, as the genetic correlations between non-suicidal self-injury and the suicidal thoughts and behaviors GWAS (r_g_ = 0.54-0.69) were, for the most part, weaker than those observed amongst the suicidal thoughts and behaviors themselves (r_g_ = 0.53-0.84) (shown in Table S5).

While genetic liability to broad sense self-harm ideation, non-fatal suicide attempt, death by suicide and depression were all associated with greater likelihood of several suicidal thoughts and behaviors, the finding that these associations even remain significant in some multi-PRS models indicates that genetic liabilities to depression, broad sense self-harm ideation, non-fatal suicide attempt and death by suicide independently contribute to variance in observed suicidal thoughts and behaviors in COGA. While previous work has also shown that predisposition to suicide attempt has a genetic component independent from depression [18,25], the current study finds that genetic liability to other suicidal thoughts and behaviors, specifically broad sense self-harm ideation and death by suicide, also make contributions that are independent from depression (i.e., both PRS independently contribute to active suicidal ideation, and broad sense self-harm ideation PRS independently contributes to suicidal ideation). These results add to evidence from epidemiological studies that have found a family history of suicidal thoughts and behaviors significantly increases risk of suicidal thoughts and behaviors independently of family history for psychiatric illness (e.g., depression) [53]. Furthermore, we find that PRS for suicidal thoughts and behaviors, particularly non-fatal suicide attempt PRS, may be better predictors of some suicidal thoughts and behaviors than PRS for depression. For example, non-fatal suicide attempt PRS was significantly associated with desire to die and attempt severity while depression PRS was not, non-fatal suicide attempt PRS showed a stronger association with active suicidal ideation than depression PRS in univariate models, and depression PRS was no longer significantly associated with persistent suicidal ideation after accounting for non-fatal suicide attempt PRS in a multi-PRS model. Thus, studies predicting suicidal thoughts and behaviors should not rely on depression PRS alone.

The death by suicide PRS was predictive of suicidal ideation, active suicidal ideation, suicide attempt and severity of suicidality in univariable models, although only the association with active suicidal ideation remained significant in the multi-PRS models. The non-suicidal self-injury PRS was only predictive of non-suicidal self-injury, suicidal ideation and suicide attempt, although none of these associations remained significant after accounting for the other associated PRS. We consider that the relatively smaller sample sizes of the death by suicide GWAS (N= 18,223, N cases = 3,413) and non-suicidal self-injury GWAS (N= 133,620, N cases= 2,845) may be contributing to the differences observed in the non-suicidal self-injury and death by suicide PRS, considering that sample size of the discovery GWAS is an important factor in determining accuracy and predictive power of PRS [49,54]. Based on the SNP-heritability, prevalence and sample sizes of these traits, we estimate our power to detect an association between the non-suicidal self-injury PRS and non-suicidal self-injury in COGA to be only 5.6%, and between the death by suicide PRS and suicide attempt in COGA to be between 40.51% and 48.99%. Additional factors beyond the smaller sample size of the GWAS may have influenced our lack of findings for the death by suicide PRS. For instance, individuals who are at the highest levels of genetic risk for death by suicide may not be adequately represented in a longitudinal study such as COGA. Furthermore, the death by suicide GWAS included individuals who died by varied violent and nonviolent means [19], and as such may reflect a highly heterogeneous set of genetic liabilities [60].

We did not observe associations between PRS and several suicidal thoughts and behaviors, specifically no PRS were significantly associated with endorsing substance use prior to attempt, requiring medical treatment after attempt, or reporting more than one attempt. Additionally, several suicidal thoughts and behaviors (e.g., desire to die, persistent suicidal ideation) were only significantly associated with one or two PRS. While it is possible that these behaviors may not have as strong of a genetic component, we also consider that the nestedness of these phenotypes may be contributing to these observations; these suicidal thoughts and behaviors specifically represent co-occurring behaviors, such that they were only asked of individuals who had endorsed suicidal ideation or suicide attempt and as such, any individual not endorsing the prerequisite variable was not included in the follow-up variables (e.g., for requiring medical treatment after an attempt, the unaffected group only included individuals who had made a suicide attempt). As a result, the “control” groups for these variables represent an already high-risk group and we consider that while genetic liability contributes to suicidal thoughts and behaviors broadly, related features of individual suicidal thoughts and behaviors may not be influenced by the PRS studied by us. It is also plausible that some features of individual suicide attempts may be situational; for example, requiring medical treatment after an attempt may reflect lethality and mode of attempt, which may be environmental in nature [61–63]. Furthermore, individuals with multiple suicide attempts only answered follow-up questions in reference to their self-defined most serious attempt. For example, an individual may have made a previous attempt after using substances, but if they had a made a different attempt they believed to be more serious in which they didn’t use a substance before the attempt, then they answered no to substance use prior to attempt. As a result, some of the behaviors co-occurring with suicide attempt that we studied may not accurately represent an individuals’ history of these behaviors over their lifetime.

In addition to small discovery GWAS sample sizes for some traits and nestedness of some variables in COGA, we acknowledge various other limitations to this study. First, COGA was ascertained for families with an extensive history of AUD, and as such is not representative of the broader population. Since individuals with substance use disorders are at higher risk for suicidal thoughts and behaviors [64,65], our sample may be enriched for these phenotypes. Second, concerns of accuracy and limited availability relating to data on family history of suicidal thoughts and behaviors prevented us from constructing models in which PRS and family history simultaneously predicted suicidal thoughts or behaviors, a method shown to improve risk prediction for other disorders [66,67]. Third, as is the case for many PRS, these PRS may be strongly associated with suicidal thoughts and behaviors at the population scale, but are not useful for individual level prediction. This is not to say PRS for suicidal thoughts and behaviors will never be useful for classification purposes, but currently they are not sufficiently powered, nor have important ethical concerns been addressed [68] for prediction at the individual or clinical level. Finally, because of the limited non-European ancestry GWAS available for suicidal thoughts and behavior phenotypes, we were only able to conduct these analyses in a subset of individuals in COGA with European ancestry, despite the diversity of the full COGA sample.

In conclusion, we find evidence that PRS computed from GWAS of non-suicidal self-injury, broad sense self-harm ideation, non-fatal suicide attempt, death by suicide and depression are significantly associated with increased risk of suicidal thoughts and behaviors in individuals in COGA. Results further corroborate that suicidal thoughts and behaviors are heterogeneous and partially genetically distinct from depression.

## Data Availability

All previously published summary statistics used in these analyses are publicly available. The non-suicidal self-injury data were derived from the UK Biobank (https://www.ukbiobank.ac.uk/) under application number 19798. The non-suicidal self-injury GWAS summary statistics generated in this study are publicly available (https://github.com/WashU-BG/nssi_gwas).

## Acknowledgements

The Collaborative Study on the Genetics of Alcoholism (COGA), Principal Investigators B. Porjesz, V. Hesselbrock, T. Foroud; Scientific Director, A. Agrawal; Translational Director, D. Dick, includes eleven different centers: University of Connecticut (V. Hesselbrock); Indiana University (H.J. Edenberg, T. Foroud, Y. Liu, M.H. Plawecki); University of Iowa Carver College of Medicine (S. Kuperman, J. Kramer); SUNY Downstate Health Sciences University (B. Porjesz, J. Meyers, C. Kamarajan, A. Pandey); Washington University in St. Louis (L. Bierut, J. Rice, K. Bucholz, A. Agrawal); University of California at San Diego (M. Schuckit); Rutgers University (J. Tischfield, R. Hart, J. Salvatore); The Children’s Hospital of Philadelphia, University of Pennsylvania (L. Almasy); Virginia Commonwealth University (D. Dick); Icahn School of Medicine at Mount Sinai (A. Goate, P. Slesinger); and Howard University (D. Scott). Other COGA collaborators include: L. Bauer (University of Connecticut); J. Nurnberger Jr., L. Wetherill, X., Xuei, D. Lai, S. O’Connor, (Indiana University); G. Chan (University of Iowa; University of Connecticut); D.B. Chorlian, J. Zhang, P. Barr, S. Kinreich, G. Pandey (SUNY Downstate); N. Mullins (Icahn School of Medicine at Mount Sinai); A. Anokhin, S. Hartz, E. Johnson, V. McCutcheon, S. Saccone (Washington University); J. Moore, Z. Pang, S. Kuo (Rutgers University); A. Merikangas (The Children’s Hospital of Philadelphia and University of Pennsylvania); F. Aliev (Virginia Commonwealth University); H. Chin and A. Parsian are the NIAAA Staff Collaborators. We continue to be inspired by our memories of Henri Begleiter and Theodore Reich, founding PI and Co-PI of COGA, and also owe a debt of gratitude to other past organizers of COGA, including Ting-Kai Li, P. Michael Conneally, Raymond Crowe, and Wendy Reich, for their critical contributions. This national collaborative study is supported by NIH Grant U10AA008401 from the National Institute on Alcohol Abuse and Alcoholism (NIAAA) and the National Institute on Drug Abuse (NIDA).

## Statement of Ethics

This research was conducted at Washington University in St. Louis under approved IRB protocol # 201906072. All participants in the COGA sample provided written informed consent.

## Author Contributions

ECJ and SMCC designed the project. SMCC performed analyses. ECJ supervised the study. All authors contributed to interpretation of results. SMCC and ECJ wrote the manuscript and all authors contributed to editing and revising the manuscript.

